# Depressive, anxiety and stress symptoms among university students from Bogotá, Colombia and electronic nicotine delivery systems (ENDS) use: Findings from a prevalence survey 2021

**DOI:** 10.1101/2024.06.18.24309072

**Authors:** María Camila García Durán, María Catalina Botero Ruge, Gustavo Perdomo Patiño, Adriana Pulido Álvarez, Manuela Lüchau, Elizabeth Borrero, Sergio Moreno

## Abstract

The use of electronic nicotine delivery systems (ENDS) has increased in recent years, particularly among young people. The association between mental health disorders and conventional cigarettes is well established. However, information on ENDS use and mental health is still emerging. This study i) determines the frequency of depression, anxiety and stress symptoms among university students in Bogotá, Colombia, ii) examines the association between sociodemographic variables and mental health symptoms, and iii) analyses whether there is an association between tobacco and/or ENDS use and depression, anxiety and/or stress. The study is cross-sectional, analytical, retrospective and based on a multistage random sample. 3850 students from 21 universities completed an online survey and were included as participants. We assessed demographic variables, tobacco use, ENDS use and symptoms of depression, anxiety and stress. 667 participants from our sample had a previous diagnosis of a mental disorder, and the most common diagnoses were any type of anxiety disorder (7.27%) or depressive disorder (6.70%). Ever use of conventional cigarettes was 63.71% and ever use of ENDS was 31%. More than 65% of the participants had mild symptoms of depression, anxiety and stress. However, between 16.4% and 19.5% of participants reported severe or very severe symptoms of these three diagnoses. Severe or very severe symptoms were more likely to be reported by women, public university students and those from higher socio-economic backgrounds. Those who reported having tried conventional cigarettes or ENDS had more severe or extremely severe symptoms than those who had never tried them. Multivariate analysis using ordinal logistic regression showed that depression, anxiety and stress levels were more severe in participants who had tried ENDS. In addition, having nuclear family members, family members or close friends with a history of nicotine use was positively associated with higher levels of depression, anxiety and stress.

## Introduction

Electronic nicotine delivery systems (ENDS), also known as electronic cigarettes (e-cigarettes) or vapers, are battery-powered devices used to inhale, smoke or ’vape’ an aerosol that usually contains flavourings and nicotine, as well as a range of other chemicals such as propylene glycol, glycerol and vegetable glycerine, and a range of metals that can be highly toxic, such as lead, chromium and nickel [1]. These devices began to be marketed in the United States around 2010, and their use has increased rapidly in recent years [2].

According to the American Academy of Paediatrics (2019), the rate of people in the United States who have tried electronic nicotine delivery systems (ENDS) at least once in their lifetime doubled between 2013 and 2016. In 2016, the overall prevalence of ever-users of electronic nicotine delivery systems (ENDS) was about 16.4%, and the prevalence of current users was 5.6%. Among non-tobacco smokers, the prevalence of ever-users of ENDS was 7.0% and the prevalence of current users was 1.5%. The prevalence of use of these devices among the tobacco smoking population showed that 54.7% had ever used ENDS, while 19.4% were current users of both tobacco and ENDS [3].

The increase in the use of ENDS may be related to the fact that these devices are perceived as less harmful than regular cigarettes [4], as a mechanism for socialising with peers due to the social rituals built around them [5], or as a smoking cessation tool. However, ENDS are controversial because of their long-term health effects [6], [7], [8] and their impact on smoking cessation is unclear in the literature [9], [10], [11], [12]. In particular, among adolescents and young adults, there is evidence of an association between e-cigarette use and increased risk of cigarette smoking initiation [13], [14], as well as increased average and frequency of cigarette consumption [15].

The link between mental health and the use of conventional cigarettes is well established. Tobacco smoking is associated with an increased risk of developing depression or anxiety [16], [17]. Adults with mental health problems have disproportionately high rates of cigarette smoking [18], [19], [20] and have a lower life expectancy than the general population as a result of this habit [21]. However, information on e-cigarette use and mental health is still emerging.

A nationally representative study of adults in the United States found that ENDS-only, cigarette-only or dual users were more likely to report ever and in the past 12 months internalising and externalising problems than those who did not report using tobacco products [22]. Similarly, other studies suggest that people with mental health problems are more likely to have tried and/or used e-cigarettes than those without mental health problems or psychological distress [23], [24], [25], [26]. The findings are similar among young adults. Studies suggest that students with a history of mental health problems are more likely to use cigarettes [27], [28], [29], and that vaping is associated with conventional smoking, cannabis use and/or alcohol use [30], [31], [32].

The relationship between depressive symptoms and ENDS use has been studied more extensively. Some cross-sectional studies in young populations suggest that ENDS use is associated with increased odds of reporting depressive symptoms and suicidal ideation [33], [34]. One longitudinal study found a bidirectional association between depressive symptoms and e-cigarette use [35]. Greater depressive symptoms at age 14 years were associated with a faster rate of e-cigarette escalation [36]. Findings in adults are similar. People who use e-cigarettes are more likely to report depression [37], and conversely, depressive symptoms have shown positive associations with ENDS use and nicotine concentration in both cross-sectional and longitudinal analyses [38], [39].

Similar associations have been found between anxiety and/or stress and ENDS use. Social anxiety has been indirectly associated with e-cigarette initiation [40], and higher anxiety symptoms have been associated with current cigarette and e-cigarette use among college students [41]. A study of a nationally representative sample of US adults reported that e-cigarette use was associated with increased levels of psychological distress [25], [42] and also reported that perceived stress was higher among ENDS users compared with non-users. Perceived stress has been associated with a higher risk of transition to current e-cigarette use among ever-users and continued use among current users [42], [43].

Colombia has the second highest use of these devices in the Andean region (16.1%), ahead of Ecuador (27.4%) and followed by Bolivia and Peru (12.8% and 12.5% respectively) [44]. ENDS are the third most used legal substance in this country, after alcohol and conventional cigarettes [45]. The prevalence of ever use of ENDS is 15.4% among high school students [46] and 16.1 % among university students, with higher prevalence rates among males than females [44]. The population aged 18-24 years has the highest rates of ever and monthly use [45]. This age group corresponds to the age of most university students. Prevalence rates of depression and anxiety among university students are higher than 30% in some regions of the country, such as Bucaramanga, Medellin and the Atlantic coast, where studies have suggested there can be an unexplored and increased risk in this population [47], [48], [49], [50].

Information on the association between ENDS consumption and mental health among university students is not available for the Colombian population. The present study aimed to a) determine the frequency of symptoms related to depression, anxiety and stress among university students in Bogotá, b) examine the association between socio-demographic variables and these mental health conditions in our population, and c) determine if there is an association between tobacco and/or e-cigarette use and depression, anxiety and/or stress. The results obtained may identify at-risk groups, inform student health sectors, and contribute to public policies and services for university students.

## Materials and Methods

### Participants and data collection

This study is part of a project entitled: "Prevalence and psychosocial factors associated with ENDS consumption in university populations of Bogota" [51]. The design of the study was cross-sectional, analytical and based on a multistage random sample. The sample size (3985) was calculated using an estimated margin of error of 5%, an estimated prevalence of ENDS use of 16.6%, an interclass correlation coefficient of 4% and a significance level of 5%. The first sampling stage consisted of the Higher Education Institutions (HEIs) registered in Bogotá, Colombia, selected proportionally to their size based on the number of students. The second stage consisted of the random selection of programmes in each of the selected HEIs. Substitutions were made in the event of no acceptance or no response from an HEI.

Students were invited to participate in an online survey hosted in the cloud research software REDCap ®[52], consisting of 64 questions related to socio-demographic data, tobacco use, ENDS use, tobacco and/or ENDS use during the pandemic, and mental health (depression, anxiety and stress). Participants were eligible if they were 18 years or older and enrolled in one of the selected programmes and universities in Bogotá. Informed consent, approved by the Ethics Committee of the Santa Fe Foundation of Bogotá (approval code CCEI-10765-2019), was provided before the questions were administered, in accordance with the Declaration of Helsinki and all relevant guidelines and ethical regulations. The survey took approximately 10-15 minutes to complete. No financial incentives were offered. Data were collected from March 2021 to November 2021. 3850 students completed the survey and were included in the study.

### Measures

#### Demographics

We used the following demographic information as control variables in the model: gender (male/female), age, socioeconomic level (low/middle/high), type of university (private/public), number of children, and whether they had worked and studied in the last 6 months. It is important to clarify that in Colombia there are seven socioeconomic levels (from 0 to 6), which we recoded into low (0-2), middle (3-4) and high (5-6) according to what is used in the country.

**Other variables of interest** included in the analyses (and their possible responses) were a) people living with the participant (both parents / mother or father / other family member / alone / friends / couple / acquaintances / other), b) if they exercised (yes / no), c) if they had ever had a mental health diagnosis (any type of bipolar disorder / depressive disorder / post-traumatic stress disorder / anxiety disorder / schizophrenia or any psychotic disorder / personality disorder / substance or alcohol abuse / eating disorder / none / other disorder not mentioned in the list); (d) whether anyone close to them used nicotine (members of the nuclear family (mother, father, siblings) / family members / couple / close friends / people they live with (housemates)

/ other close person).

**Tobacco use:** Was assessed by the question "Have you ever tried smoking a cigarette in your life, even just once?" (yes/no).

**ENDS use:** Was measured by the question "Have you ever used an ENDS (such as electronic cigarettes or vapers)?" (yes/no).

#### DASS-21

The Depression, Anxiety, and Stress Scale (DASS-21) is a 21-item scale used to screen for depression, anxiety, and stress. This scale has been validated in Colombia and shows good internal consistency (0.92-0.93)[53]. Each subscale is assessed through 7 items. Participants are asked to score the questions from 0 (did not apply to me at all) to 3 (applied to me very much or most of the time); thus, every subscale has a maximum score of 21. Higher scores represented greater levels of the condition. The cut-off points used to determine the severity of depression were 5-6 (mild), 7-10 (moderate), 11-13 (severe), and 14 or more (extremely severe); to determine the severity of anxiety were 4 (mild), 5-7 (moderate), 8-9 (severe) and 10 or more (extremely severe); and to determine the severity of stress were 8-9 (mild), 10-12 (moderate), 13-16 (severe) and 17 or more (extremely severe).

### Statistical analysis

Descriptive analysis was performed using measures of central tendency and dispersion for the quantitative variables and absolute and relative frequencies for the qualitative variables, stratified by level of depression according to the DASS-21 scale.

The possible factors associated with the different levels of depression, anxiety and stress were assessed using an ordinal logistic regression model. The candidate variables for the model were based on the literature review and their possible contextual plausibility. Multivariate models were estimated, considering first-order interactions to assess potential confounders, and a reduced model was constructed based on the Furnival-Wilson algorithm, reporting the logarithm of likelihood, Akaike’s information criterion, and Bayesian information criterion. Adjusted odds ratios (aOR) were reported as measures of association with their corresponding confidence intervals. The goodness of fit of the model and its underlying assumptions were assessed using a linearity test, the parallelism or proportional odds test, collinearity using the VIF and analysis of deviance residuals. The a priori significance level was set at 5%. The statistical analysis was performed in Stata16MP.

## Results

A total of 3850 participants were included in this study. The demographic characteristics of the population and some other variables of interest are described in Table 1. Most of our sample (61%) were women, of the total sample 3527 (92%) were between 18 and 26 years old and 93% were studying in a private university. More than half of the participants belonged to a middle socio-economic level (53%), followed by a low socio-economic level (37.16%). Of the total sample, over 70% of the participants worked and studied at the same time in the last 6 months, and almost the same percentage reported not having any children. Most participants reported living with their parents and/or in a couple and more than half of the students reported doing some kind of physical activity (54.1%). Finally, participants reported that their family members (31.9%), close friends (30.13%) and members of their nuclear family (28.16%) used nicotine.

**Table 1.** Demographic characteristics of the study population.

| Variable |  | N (%) |
| --- | --- | --- |
|  |  | N= 3850 |
| **Sociodemographic** |  |  |
| **Sex** | **Male** | **1513 (39%)** |
|  | **Female** | **2337 (61%)** |
| **Age group** | **Between 18-26** | **3527 (91.61%)** |
|  | **Between 27-60** | **322 (8.36%)** |
|  | **Missing data** | **1 (0.03%)** |
| **Socioeconomic Level** | **Low (0-1-2)** | **1431 (37.16%)** |
|  | **Medium (3-4)** | **2041 (53.01%)** |
|  | **High (5-6)** | **334 (8.68%)** |
|  | **Missing data** | **44 (1.14%)** |
| **Type of university** | **Public** | **276 (7.17%)** |
|  | **Private** | **3574 (92.83%)** |
| **Worked and studied during the past** | **No** | **1132 (29.40%)** |
|  |  | N= 3850 |
| 6 months | Yes | 2710 (70.39%) |
|  | Missing data | 8 (0.21%) |
| Number of children | None | 2684 (70.32%) |
|  | 1 child | 606 (15.88%) |
|  | 2 children | 395 (10.35%) |
|  | 3 or more children | 132 (3.46%) |
| Other variables of interest |  |  |
| People who live with the participant | Both parents | 1203 (31.25%) |
|  | Mother or father | 952 (24.73%) |
|  | Another family member | 728 (18.91%) |
|  | Alone | 267 (6.94%) |
|  | Friends | 67 (1.74%) |
|  | Couple | 1028 (26.70%) |
|  | Acquaintances | 23 (0.60%) |
|  | Other | 267 (6.94%) |
| Exercise (physical activity) | No | 1759 (45.69%) |
|  | Yes | 2083 (54.10%) |
|  | Missing data | 8 (0.21%) |
| Previous mental health diagnosis | Any type of depressive disorder | 258 (6.70%) |
|  | Any type of bipolar disorder | 14 (0.36%) |
|  | Postraumatic stress disorder | 24 (0.62%) |
|  |  | N= 3850 |
|  | Any type of anxiety disorder | 280 (7.27%) |
|  | Schizophrenia or any type of psychotic disorder | 6 (0.16%) |
|  | Personality disorder | 16 (0.42%) |
|  | Abuse or dependence of psychoactive substances | 14 (0.36%) |
|  | Eating disorder | 55 (1.43%) |
| Close people who consume nicotine | Members of the nuclear family | 1084 (28.16%) |
|  | Other family members | 1228 (31.90%) |
|  | Couple | 307 (7.97%) |
|  | Close friends | 1160 (30.13%) |
|  | Housemates | 82 (2.13%) |
|  | Other close person | 489 (12.70%) |
| ENDS and tobacco use |  |  |
| Have tried conventional cigarette | Yes | 2453 (63.71%) |
|  | No | 1389 (36.08%) |
|  | Missing data | 8 (0.21%) |
| Have tried ENDS | Yes | 1187 (30.83%) |
|  | No | 2630 (68.31%) |
|  | Missing data | 33 (0.86%) |

667 participants had a previous diagnosis of any mental disorder, the most common being any type of anxiety disorder (7.27%) and any type of depressive disorder (6.70%). Most participants have tried conventional cigarettes (63.71%). On the contrary, only 31% of them have ever used ENDS.

Table 2 shows the levels of depressive, anxiety and stress symptoms in the study population by sex, type of university and socioeconomic level, as well as by ever using conventional cigarettes or ENDS.

**Table 2.** Levels of depression, anxiety and stress symptoms stratified by sociodemographic variables and tobacco/ENDS use.

|  |  | Depression |  |  |  | Anxiety |  |  |  | Stress |  |  |  |
| --- | --- | --- | --- | --- | --- | --- | --- | --- | --- | --- | --- | --- | --- |
|  |  | Mild | Moderate | Severe | Extremely severe | Mild | Moderate | Severe | Extremely severe | Mild | Moderate | Severe | Extremely severe |
| General |  | 2656<br>(69.0%) | 510<br>(13.2%) | 278 (7.2%) | 406 (10.5%) | 2504<br>(65.0%) | 595<br>(15.5%) | 234<br>(6.1%) | 517 (13.4%) | 2782<br>(72.3%) | 437<br>(11.4%) | 425<br>(11.0%) | 206 (5.4%) |
| Sex | Male | 1102<br>(72.8%) | 169<br>(11.2%) | 112 (7.4%) | 130 (8.6%) | 1080<br>(71.4%) | 201<br>(13.3%) | 77 (5.1%) | 155 (10.2%) | 1170<br>(77.3%) | 155<br>(10.2%) | 127 (8.4%) | 61 (4.0%) |
|  | Female | 1554<br>(66.5%) | 341<br>(14.6%) | 166 (7.1%) | 276 (11.8%) | 1424<br>(60.9%) | 394<br>(16.9%) | 157<br>(6.7%) | 362 (15.5%) | 1612<br>(69.0%) | 282<br>(12.1%) | 298<br>(12.8%) | 145 (6.2%) |
| Type of university | Public | 153 (55.4%) | 37 (13.4%) | 28 (10.1%) | 58 (21.0%) | 159 (57.6%) | 45 (16.3%) | 11 (4.0%) | 61 (22.1%) | 175 (63.4%) | 32 (11.6%) | 40 (14.5%) | 29 (10.5%) |
|  | Private | 2503<br>(70.0%) | 473<br>(13.2%) | 250 (7.0%) | 348 (9.7%) | 2345<br>(65.6%) | 550<br>(15.4%) | 223<br>(6.2%) | 456 (12.8%) | 2607<br>(72.9%) | 405<br>(11.3%) | 385<br>(10.8%) | 177 (5.0%) |
| Socioeconomic level | Low | 1036<br>(72.4%) | 175<br>(12.2%) | 93 (6.5%) | 127 (8.9%) | 982 (68.6%) | 183<br>(12.8%) | 82 (5.7%) | 184 (12.9%) | 1087<br>(76.0%) | 132 (9.2%) | 150<br>(10.5%) | 62 (4.3%) |
|  | Middle | 1398<br>(68.5%) | 274<br>(13.4%) | 139 (6.8%) | 230 (11.3%) | 1309<br>(64.1%) | 339<br>(16.6%) | 129<br>(6.3%) | 264 (12.9%) | 1460<br>(71.5%) | 243<br>(11.9%) | 217<br>(10.6%) | 121 (5.9%) |
|  | High | 197 (59.0%) | 53 (15.9%) | 42 (12.6%) | 42 (12.6%) | 186 (55.7%) | 64 (19.2%) | 23 (6.9%) | 61 (18.3%) | 201 (60.2%) | 60 (18.0%) | 55 (16.5%) | 18 (5.4%) |
|  | Missing | 25 (56.8%) | 8 (18.2%) | 4 (9.1%) | 7 (15.9%) | 27 (61.4%) | 9 (20.5%) | 0 (0.0%) | 8 (18.2%) | 34 (77.3%) | 2 (4.5%) | 3 (6.8%) | 5 (11.4%) |
| Having tried conventional cigarette | Yes | 1653<br>(67.4%) | 326<br>(13.3%) | 195 (7.9%) | 279 (11.4%) | 1567<br>(63.9%) | 389<br>(15.9%) | 145<br>(5.9%) | 352 (14.3%) | 1732<br>(70.6%) | 292<br>(11.9%) | 290<br>(11.8%) | 139 (5.7%) |
|  | No | 995 (71.6%) | 184<br>(13.2%) | 83 (6.0%) | 127 (9.1%) | 929 (66.9%) | 206<br>(14.8%) | 89 (6.4%) | 165 (11.9%) | 1042<br>(75.0%) | 145<br>(10.4%) | 135 (9.7%) | 67 (4.8%) |
|  | Missing | 8 (100%) | 0 (0.0%) | 0 (0.0%) | 0 (0.0%) | 8 (100%) | 0 (0.0%) | 0 (0.0%) | 0 (0.0%) | 8 (100%) | 0 (0.0%) | 0 (0.0%) | 0 (0.0%) |
| Having tried ENDS | Yes | 690 (58.1%) | 191<br>(16.1%) | 122<br>(10.3%) | 184 (15.5%) | 670 (56.4%) | 226<br>(19.0%) | 75 (6.3%) | 216 (18.2%) | 752 (63.4%) | 162<br>(13.6%) | 178<br>(15.0%) | 95 (8.0%) |
|  | No | 1939<br>(73.7%) | 316<br>(12.0%) | 153 (5.8%) | 222 (8.4%) | 1809<br>(68.8%) | 364<br>(13.8%) | 159<br>(6.0%) | 298 (11.3%) | 2003<br>(76.2%) | 273<br>(10.4%) | 243 (9.2%) | 111 (4.2%) |
|  | Missing | 27 (81.8%) | 3 (9.1%) | 3 (9.1%) | 0 (0.0%) | 25 (75.8%) | 5 (15.2%) | 0 (0.0%) | 3 (9.1%) | 27 (81.8%) | 2 (6.1%) | 4 (12.1%) | 0 (0.0%) |

Over 65% of participants were classified as having mild symptoms. For depression, 69% of the sample reported mild symptoms, 13.2% reported moderate symptoms and 17.7% reported severe (7.2%) or extremely severe (10.5%) symptoms. For anxiety, 65% of participants reported no or mild symptoms, 15.5% reported moderate symptoms and 19.5% reported severe (6.1%) or extremely severe (13.4%) symptoms. This pattern was relatively similar for stress. Most of the sample had no or mild symptoms (72.3%), 11.4% of them were in the moderate category and 16.4% were in the severe (11.0%) or extremely severe (5.4%) category. Severe or extremely severe symptoms of depression, anxiety and stress were more likely to be reported by women, participants attending public universities and those from higher socioeconomic levels. Those who reported having tried conventional cigarettes or ENDS had more severe or extremely severe symptoms than those who had never tried them.

Table 3 shows the multivariate analysis by ordinal logistic regression of socio-demographic and tobacco/ENDS use variables associated with depression, anxiety and stress. Depression was more severe in participants who had tried ENDS (OR: 1.35; 95% CI: 1.15-1.59). Higher levels of depression were found in women (OR: 1.39; 95% CI: 1.19-1.62), students from public universities (OR: 1.53; 95% CI: 1.19-1.98), participants who reported a previous diagnosis of any type of depressive disorder (OR: 4.80; 95% CI: 3.63-6.34) or any type of anxiety disorder (OR: 2.92; 95% CI: 2.23-3.83). Having members of the nuclear family (OR: 1.23; 95% CI: 1.05-1.44), family members (OR: 1.35; 95% CI: 1.15-1.57), or close friends (OR: 1.28; 95% CI: 1.09-1.50) with a history of nicotine use was positively associated with higher levels of depression. Participants with a child (OR: 0.62; 95% CI: 0.49-0.79), who lived in a couple (OR: 0.66; 95% CI: 0.53-0.82), or who reported studying and working at the same time in the last 6 months (OR: 0.82; 95% CI: 0.70-0.96) or doing physical activity (OR: 0.82; 95% CI: 0.71-0.95) were less likely to have worse depression.

**Table 3.**
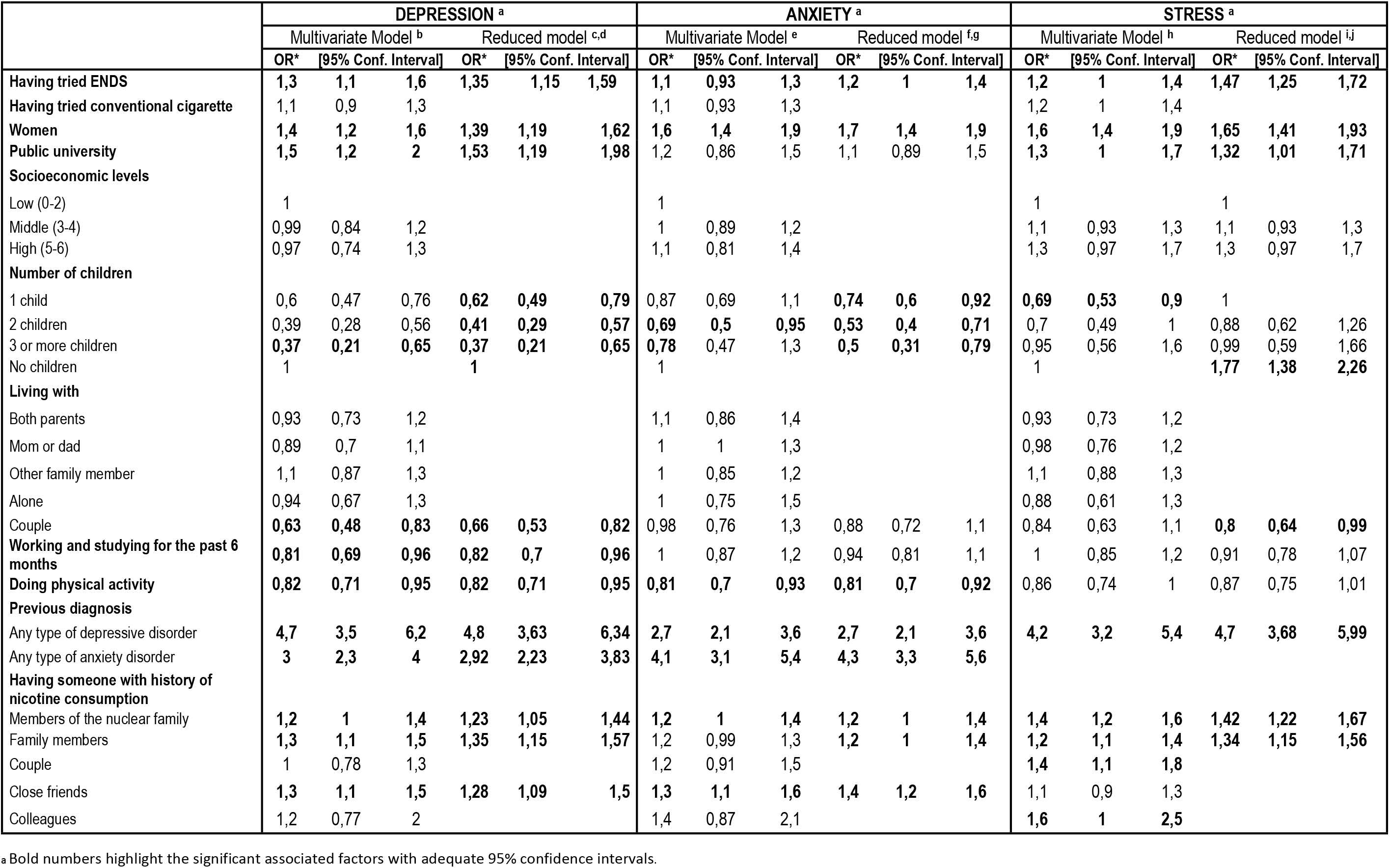
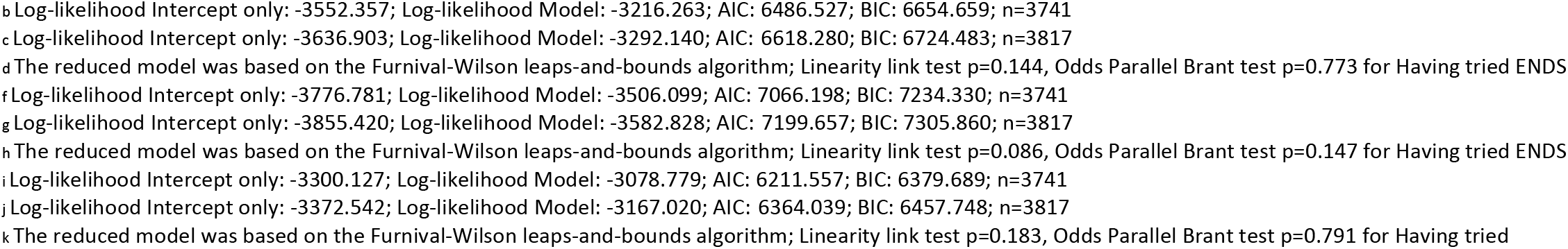
Multivariate analysis of the sociodemographic and use of tobacco/ENDS variables associated with depression, anxiety and stress.

Anxiety levels were higher in women compared to men (OR: 1.7; 95% CI: 1.4-1.9) and in people who had tried ENDS (OR: 1.2; 95% CI: 1-1.4). Participants with a history of depressive disorders (OR: 2.7; 95% CI: 2.1-3.6), anxiety disorders of any type (OR: 4.3; 95% CI: 3.3-5.6), or those who reported having close relatives (OR: 1.2; 95% CI: 1-1.4), family members (OR: 1.2; 95% CI: 1-1.4), and friends (OR: 1.4; 95% CI: 1.2-1.6) with a history of nicotine use were more likely to have higher levels of anxiety. On the other hand, people who reported being parents of three or more children (OR: 0.5; 95% CI: 0.31-0.79) and those who did some kind of physical activity (OR: 0.81; 95% CI: 0.7-0.92) had lower levels of anxiety.

A previous diagnosis of any type of depressive disorder increased the likelihood of having higher levels of stress by 4.7 times (OR: 4.7; 95% CI: 3.6-5.9). Women (OR: 1.65; 95% CI: 1.4-1.9), people who had tried ENDS in their lifetime (OR: 1.47; 95% CI: 1.25-1.72) and students at public universities (OR: 1.32; 95% CI: 1.0-1.7) had a higher risk of having worse stress levels. This result is similar for people who had members of the nuclear family (OR: 1.4; 95% CI: 1.2-1.6), as well as for family members (OR: 1.3; 95% CI: 1.1-1.5) with a history of nicotine consumption, or for people without children (OR: 1.77; 95% CI: 1.3-2.2). Finally, participants who lived with a couple had fewer stress symptoms (OR: 0.8; 95% CI: 0.6-0.9).

## Discussion

This study with university students from Bogotá revealed high frequency of symptoms of depression, anxiety and stress in the population, with higher levels of mental health symptoms in people with a previous diagnose of depression or anxiety. This finding is similar to those reported in other small studies with participants from different regions of Colombia [48], [49], [50], [54] but significantly different from the prevalence reported for these age groups in the latest National Mental Health Survey of Colombia (NMHS, 2015) [55]. According to the NMHS, the prevalence of a depressive disorder in the last 30 days is 0.5% in adults and 1.1% for an anxiety disorder [55].

There are several factors that could explain this difference. First, the present study assesses symptoms of depression and anxiety, whereas the NMHS assesses the prevalence of diagnosed disorders. In addition, the prevalence of mood disorders among students has increased over the years [56], [57]. The COVID-19 pandemic has also had a strong impact on mental health, not only in students but also in the general population. Fear of the disease, isolation and other health-related situations experienced in the year 2020 favoured the increase in the prevalence of mental disorders worldwide [58], [59].

The Colombian NMHS describes that affective and anxiety disorders are more common in women [55], which is consistent with the findings of the present study. Both the NMHS and studies aimed at identifying the association between mental health and socio-demographic factors [60] have found a relationship with socioeconomic level of individuals. Results suggest that people with greater socioeconomic vulnerability in their households are more likely to have mental health symptoms. In the case of this study, no relationship was found between socioeconomic level and mental health symptoms.

However, we did find a relationship between the type of university the participants attended and the presence of stress and depressive symptoms. We found that students from public institutions were more likely to report more depressive and stress symptoms than those from private universities. Evidence from schools in the United Kingdom has shown that there is a difference in the mental health of students when comparing public and private schools, attributing the differences to contextual factors and to the amount of resources available for the school to invest in the student’s wellbeing and educational processes [61]. Although these data come from research in schools, it is possible that these hypotheses could explain the difference found in the present study, given that in Colombia public universities tend to have fewer resources and are highly competitive environments, being much affordable than private institutions. However, it would be important to study this phenomenon further in university contexts to confirm this hypothesis.

Working and studying at the same time was found to have an association with emotional symptoms. It could be expected that an increase in the burden of combining academic and work responsibilities would increase distress, however, the findings of this study showed the opposite. Having studied and worked simultaneously in the last six months showed a decrease in the likelihood of reporting symptoms of depression, and no significant relationship was found between this variable and symptoms of stress or anxiety. A possible explanation for this finding is that working and studying is not seen as an additional burden, but as a source of economic stability, which in turn can reduce negative emotional symptoms.

Similarly, having children could be seen as a possible additional stressor, nevertheless our model suggests that it reduces the likelihood of reporting symptoms of anxiety and depression. This is consistent with the findings of Lee et al. (2022), who showed that during the pandemic, adults living with children at home had lower rates of isolation-related distress in terms of depression and anxiety [62]. These authors suggest that this may be due to a greater sense of purpose in parenting, as well as the development of social relationships with and through children (such as participation in parenting groups), both of which may contribute to better mental health [62]. The findings of these authors may be a possible explanation to the positive effect of living with a couple on depression and stress as well.

The effect of having smokers in the immediate environment also showed a relationship with the mental health of the participants. In 2021, Lee and Kim published an article discussing how adolescent exposure to second hand smoke could increase the risk of mental health problems such as stress, depression, and suicidal ideation [63]. The results of our study showed that nicotine use by family members, other relatives or close friends increased the likelihood of having more symptoms of depression and anxiety, and that use by relatives increased the likelihood of having symptoms of stress. This may be due to both social and biological factors. On the one hand, nicotine consumption is seen as a factor affecting the ability to lead a healthy lifestyle, which has a significant impact on people’s physical and mental health, and nicotine environments can be stressful for people [64]. Secondly, nicotine exposure has been found to affect the dopaminergic pathways in the brain, which may ultimately affect people’s emotional and behavioural regulation [65]. Opposite to this, doing physical activity – a key element of a healthy lifestyle - was found to be protective for symptoms of depression and anxiety. This result has been repetitively found in multiple studies, including some systematic reviews that have shown there is a relationship between increased physical activity and the reduction of emotional symptoms associated with depression and anxiety [66]. Specifically, exercise has been found to have a positive impact on mental health due to different factors, such as a reduction in muscle tension as well as an increase in the production of certain neurochemicals associated with mood. In addition, focusing attention on physical activity has been shown to help people withdraw their attention from negative thought cycles that are associated with the presence of symptoms of depression and anxiety [67].

Finally, our results showed that having tried ENDS increases the likelihood of experiencing symptoms of depression, anxiety and stress. Interestingly, we did not find an association between having tried conventional cigarettes and any mental health symptom. Out findings contribute to other studies that are starting to show a bidirectional association between nicotine use and mental health symptoms [35], [68]. These findings may be partially explained by the perceived desirable effects of tobacco use, as many people see it as a strategy to reduce stress and emotional discomfort. However, these effects are due to the immediate physiological response produced by nicotine and other chemicals in the brain that stimulate the brain’s reward system. This brief sensation makes smoking desirable for people with various mental health conditions, as it increases the feeling of control over discomfort. However, the effect is short-lived and, in the long term, smoking can make symptoms worse [68], [69]. This can be seen, for example, in people with anxiety or stress. Initially, nicotine produces a feeling of relaxation and calm that helps to relieve anxiety symptoms, but as this effect wears off, withdrawal symptoms can worsen the symptoms of anxiety and discomfort [70]. Not only a mental health diagnosis, but also a negative perception of the impact of the pandemic on mental health has also been associated with a higher likelihood of ENDS use among Colombian university students [51].

In conclusion, our study aimed to investigate the association between the use of electronic nicotine delivery systems (ENDS) and mental health among university students in Colombia, a topic that has been understudied in the local context. Through this study, we aimed to shed light on the frequency of depression, anxiety and stress symptoms among university students, analyse the interplay between socio-demographic variables and mental health conditions, and examine the potential link between ENDS use and mental well-being. The results of our research provide valuable insights into understanding the complex relationship between ENDS use and mental health outcomes.

This study has implications for public health policy and interventions targeting university students. Future research should delve deeper into the underlying mechanisms driving the observed associations, allowing for the design of more targeted and effective interventions. Overall, our findings highlight the importance of addressing mental health issues and nicotine use together to create healthier and more resilient university environments. Furthermore, further research should focus on the need for comprehensive tobacco control policies and targeted interventions to mitigate the potential adverse effects on mental health. The identification of socioeconomic vulnerability as a factor contributing to ENDS use and associated symptoms highlights the importance of tailored strategies to support vulnerable populations and promote mental well-being. Among the strengths of this study, we highlight that the study sample included students from different universities, based on a probabilistic sampling, with the aim of obtaining inferential results and representing the results of the study. In addition, this is the first study of its kind in Colombia that analyses the relationship between ENDS consumption and mental health symptoms. Furthermore, as ENDS consumption is a relatively recent phenomenon, the available information on the characteristics of ENDS consumption is limited. This type of study is of great value in generating evidence that provides an overview of the use of these devices and assists decision-makers in designing public health policies that ensure the well-being of the general population.

Despite its strengths, our study has some limitations. First, due to the cross-sectional design of the study, it is not possible to establish a causal relationship between the demographic and consumption variables and the presence of symptoms of depression, anxiety and stress. In this case, we can only show associations between the variables. For this reason, it would be important to carry out studies with other designs in order to extend the available information on the relationship between socio-demographic factors, ENDS consumption and mental health symptoms. Secondly, although a probability sample was constructed with the intention of making the conclusions generalisable, it is important to mention that there was a greater participation of students from private universities during the process, so there is an overrepresentation of this population. In addition, the data collected were self-reported, which may affect the reliability of the information, especially when reporting mental health symptoms associated with social stigma, potentially leading to social desirability bias.

## Data Availability

Data is not available due to ethical restrictions. Due to the sensitive nature of the research supporting data is not available.

## Acknowledgements

We would like to acknowledge and thank Sandra Liliana Osses R. and Diana Carolina Maldonado V., who were part of our research team and contributed significantly to the findings of this project.

